# Early adolescent substance use patterns and associated factors: A longitudinal analysis of the Adolescent Brain Cognitive Development Study

**DOI:** 10.1101/2025.06.24.25330203

**Authors:** M.I. Harwood, D.F. Otero-Leon, E.J. Stringfellow, A. Deutsch, M.S. Jalali, H. Dong

## Abstract

**Background:** Early substance use is associated with long-term negative health and behavioral outcomes. While extensive research has examined predictors of substance use in adolescents and adults, relatively little is known about the risk factors that drive substance use in younger children. Furthermore, most studies focus on individual substances in isolation, rather than exploring the use of multiple substances. This study aims to identify demographic, family, and peer-related factors associated with alcohol, nicotine, cannabis, and multiple substance use in children aged from 9 to 13.

**Methods:** We analyzed longitudinal data from 11,868 children enrolled in the Adolescent Brain and Cognitive Development (ABCD) Study (version 5.1), incorporating observations collected from baseline, annual follow-ups, and mid-year phone interviews. Substance use outcomes were categorized into five mutually exclusive groups: no use, alcohol only, nicotine only, cannabis only, and use of two or more substances. We applied a generalized estimating equation model to assess the associations between substance use outcomes and demographic, family, and peer characteristics.

**Results:** Peer substance use was consistently associated with all categories of child substance use, particularly for the use of two or more substances. Parental drug use, permissiveness toward alcohol, and lower parental education were also linked to increased risk. Older age was associated with alcohol, nicotine, and multiple substance use.

**Conclusions:** Findings highlight the importance of peer and family environments in shaping early substance use behaviors. Early prevention efforts should consider these factors to effectively target at-risk children.

## INTRODUCTION

Substance use has long been recognized as a critical public health concern, with substantial evidence linking early initiation to a wide range of adverse outcomes, including increased risk of substance dependence, mental health disorders, cognitive impairment, academic underachievement, and involvement in the criminal justice system. While prevention efforts often focus on adolescents, evidence suggests that substance use behaviors and risk factors could develop earlier. For example, by age 12, 6.8% of youth report alcohol use, a figure that rises to 53.2% by age 17. Similarly, cannabis use increases from 1.3% at age 12 to 35.9% age 17.

Nicotine use is also notable, with 7.4% of middle schoolers and 25.3% of high schoolers reporting use. Despite this, relatively little is known about the substance use patterns and associated risk factors in early adolescence, representing a significant knowledge gap in both research and policy.

Identifying children at risk for substance use is crucial for timely and effective intervention. The early adolescent period is marked by rapid developmental changes in the brain, particularly in areas involved in decision-making, reward processing, and impulse control. Exposure to substances during this critical developmental window may have amplified long-term consequences. Moreover, the behavioral and environmental factors that contribute to substance use in early adolescence may differ from those seen in older teens or adults. For instance, while peer pressure and social contexts are important at all ages, family dynamics and early modeling of substance-related behaviors may play an even more central role in younger children. Therefore, identifying the early risk factors for substance use before adolescence is essential to shaping prevention efforts that are developmentally appropriate and maximally effective.

Current literature on youth substance use focuses on individual substance such as alcohol, nicotine, cannabis, or other illicit drugs. However, recent trends suggest that multiple substance use is increasingly common in youth and may follow a distinct, and potentially more concerning, developmental trajectory compared to those who initiate use with single substance. For example, adolescents who engage in binge drinking are at heightened risk for both subsequent alcohol and drug dependence. Despite these concerns, limited research has examined the prevalences and associated risk factors of multiple substance use, especially during late childhood. A comprehensive understanding of early multiple substance use requires identifying both shared and substance-specific risk factors across different substance use patterns. To address these gaps, we leverage data from the Adolescent Brain and Cognitive Development (ABCD) Study, the largest long-term study of brain development and child health in the United States. The ABCD Study follows a nationally representative cohort of 11,686 children recruited at ages 9–10 and tracks their development through early adulthood.

In this study, we examine the demographic, family, and peer characteristics associated with the use of alcohol, nicotine, cannabis, and two or more of these substances in children. The goal of this research is to provide a clearer understanding of the early risk environment for substance use in children, with the broader aim of informing early prevention and intervention strategies. By identifying the characteristics that differentiate children who initiate substance use, particularly those who use multiple substances, we hope to contribute actionable insights that can guide educators, clinicians, and public health practitioners in reducing the burden of substance use from an early age.

## METHODS

We utilized data the Adolescent Brain and Cognitive Development (ABCD) Study, the largest long-term study of brain development and child health in the United States. The ABCD Study has 11,868 children enrolled, and as of the 5.1 version, they are followed from 2018 to 2023. Additionally, the substance use module consists of baseline information, 4 yearly follow-up questionnaires, and 4 mid-year follow-ups.

The primary outcomes of interest were substance use, defined in five mutually exclusive categories: no use, alcohol use only, nicotine use only, cannabis use only, and multi substance use (defined as the use of two or three substances among alcohol, nicotine, and cannabis within the same year). Children who reported no use of any of these substances served as the reference group. Alcohol use was defined as the consumption of at least one full drink of beer, wine, or liquor (rum, vodka, gin, whiskey). Nicotine use included the use of any product containing nicotine, such as cigarettes, e-cigarettes, chewing tobacco, cigars, hookah, tobacco pipes, and nicotine patches. Cannabis use included any products containing cannabis, including blunts, vapes, edibles, oils, drinks, and CBD products. This information was gathered on two separate occasions during the year: the yearly follow-up that consists of multiple questionnaires asking them about their use in the past year, and a mid-year phone call follow-up that ask the participants to recall their use in the past 6 months.

Given the relatively high prevalence of alcohol, nicotine, and cannabis use in the ABCD cohort, our analysis focused on these three substances. We excluded 0.2% of total observations where participants reported using only other substances, due to their low frequency. Additionally, baseline assessments differed from follow-up waves, as participants were asked whether they had ever used each substance at any point in time prior to the survey. To ensure consistency and temporal relevance, we restricted our analysis to data from the four yearly follow-up assessments and the four mid-year follow-ups to examine risk factors associated with different patterns of substance use.

To examine factors associated with different substance use patterns, we included a wide range of demographic, family-related, and peer-related information. Demographic information was collected at baseline and included sex (female; male), race (White; Black; Other), and age. Age was updated at each yearly follow-up visit and treated as a time-varying variable. Family-related information was reported by parents or guardians and updated annually. We considered factors including parental marital status (married; unmarried), household income (lower or equal to $50,000; between $50,000 and $100,000; higher than $100,000; not reported), parental education (high school diploma or less; bachelor’s degree; post-graduate degree). We also included a binary variable indicating whether having household alcohol rules, assessed by whether the child was allowed to consume alcohol at home (yes vs. no). Parental substance use was categorized separately for alcohol and other drugs, based on whether at least one of the parents reported using alcohol or other drugs (yes vs. no). Peer-related information was reported by the child and updated annually. We included whether the child reported that their peers consumed alcohol, nicotine, marijuana, inhalants, or other substances (yes vs. no). Additionally, whether the child reported that peers sold substances was included (yes vs. no).

We first examined the baseline characteristics of the included study participants overall as well as stratified by whether participants ever used alcohol, nicotine, or cannabis during the study period. They were further compared using sex, race, income, parents education, parents marital status, parents alcohol and drug use, alcohol allowed at home, peers substance use, and peers that sell substances for categorical variables and age for continuous variables. Next, we used bivariable and multivariable generalized estimating equations (GEE) to assess factors associated with the use of different substances. The outcome was modeled as a nominal variable with five categories using a log link function. The model specified an exchangeable working correlation structure to account for the potential correlations among repeated measurements within individuals over time. The GEE model was implemented in R using the *nomLORgee* function from the *multgee* package (version 1.9.0). All p-values were two-sided with significant level as 0.05.

## RESULTS

After sample restrictions, we included a total of 11,303 participants in our analysis. Table 1 presents the demographic characteristics, family- and peer-related factors at baseline. The study has mostly White participants (74.3%), followed by Black participants (16.8%), and 52.1% of them are male. During the study period, 945 (8.4%) of the participants have use any substance in their lifetime, where 74.8% are White and 52.9% are male.

**Table 1.**
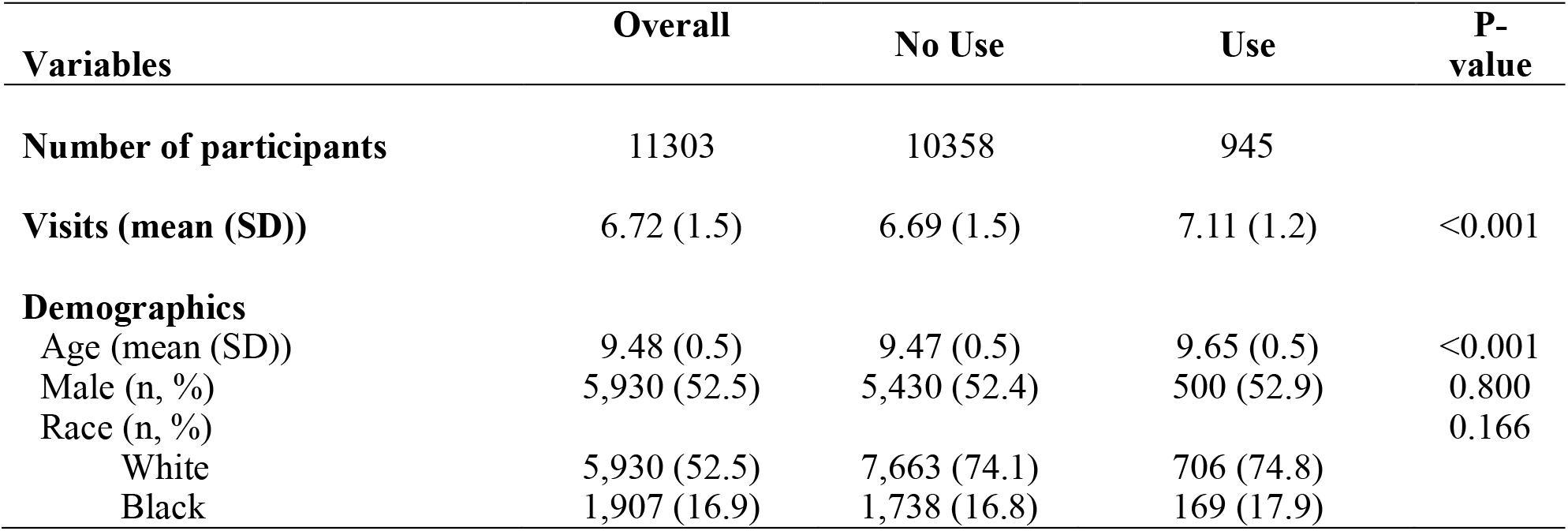
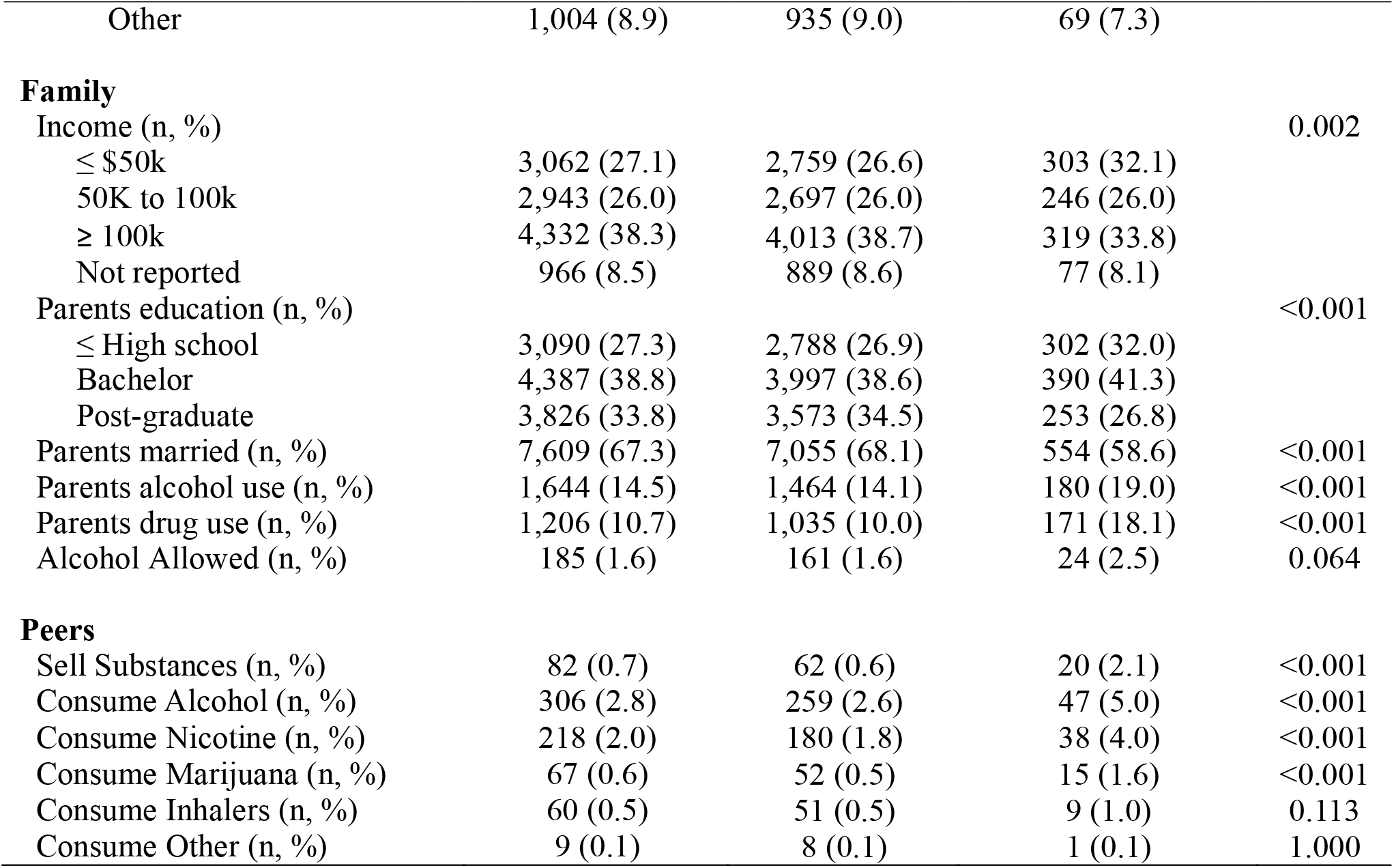
Baseline demographics, family-related, and peer-related characteristics for 11,303 youths in the ABCD Study from 2018-2023, overall and stratified by ever reported using alcohol, nicotine, and cannabis during the study period.

Overall, most families have a yearly income higher than $100,000 (38.3%), where most children who have used substances belong to these families (33.8%), followed by children from family with income under $50,000 (32.1%). Similarly, 33.8% of the parents/guardian’s education includes post-graduate studies overall, while this percentage decreases to 26.8% for children that have used substances. Similar differences are seen in other family characteristics, such as lower percentage of married parents (68.1% vs. 58.6%), higher parents’ alcohol (14.1% vs. 19.0%), and parents’ drug use (10.0% to 18.1%).

The results of the multivariable GEE models studying the characteristics that increase the risk of using alcohol, nicotine, cannabis, and the use of two or three of these substances are presented in Figure 1. Older age was associated with significantly increased odds of alcohol use (adjusted odds ratio [AOR]: xx, 95% confidence interval [CI]: 1.37,1.74). Compared to White, children who were Black (AOR: xx, 95% CI: 0.31,0.92) or Other race (AOR: xx, 95% CI: 0.14,0.65) had lower likelihood of alcohol use. Parental allowance of alcohol consumption (AOR: xx, 95% CI: 5.05, 10.14) was strongly associated with increased alcohol use. Peer factors were also significant, where having peers who consumed alcohol (AOR= (4.54, 10.28)) increased the odds of alcohol use. Finally, parental drug use (AOR= (1.02, 2.73) was also significantly associated.

**Figure 1.**
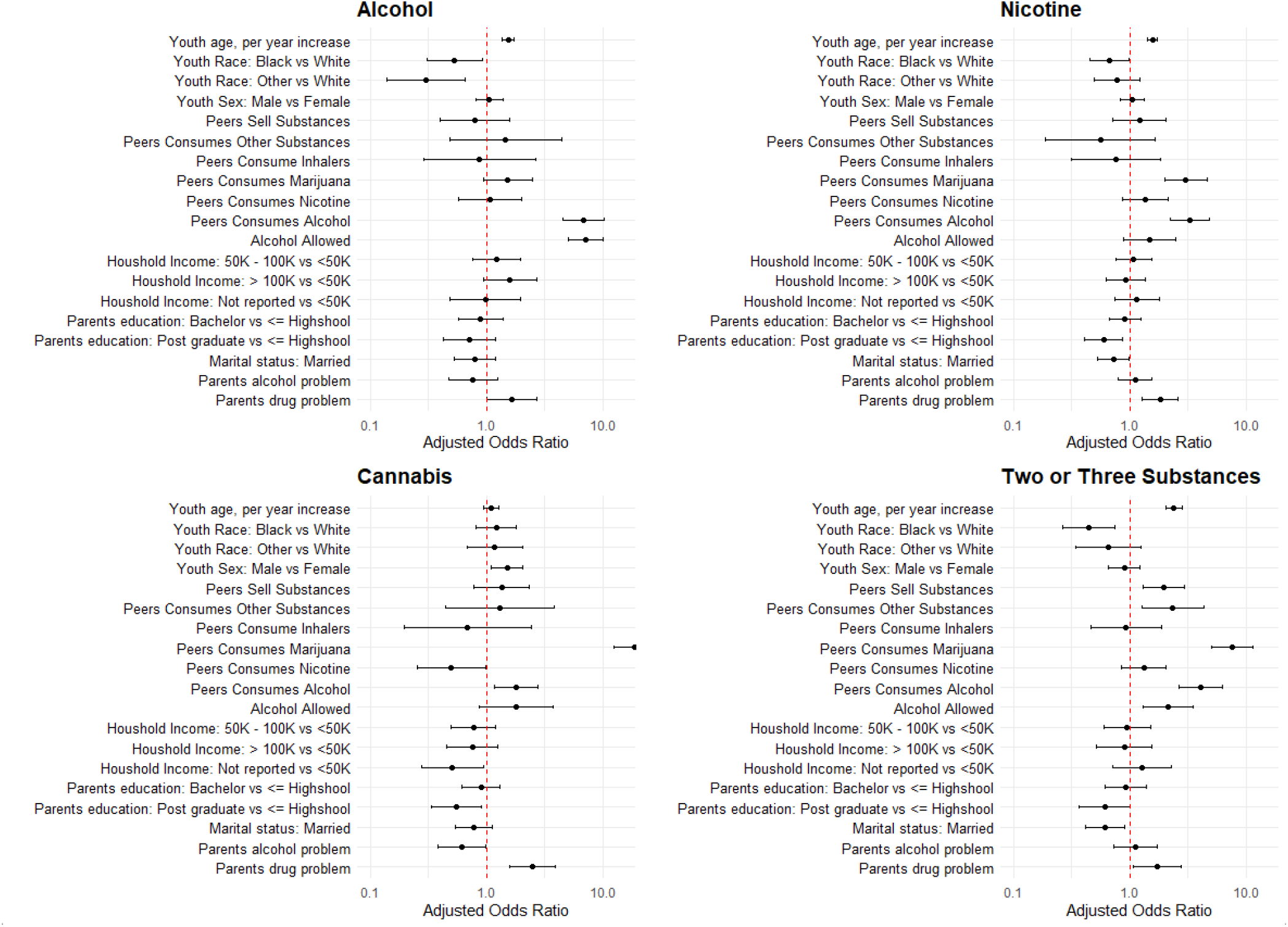
Demographics, family-related, and peer-related characteristics associated with the use of different substances in xx youths in the ABCD study from 20xx-20xx.

Nicotine use had similar significant factors as alcohol use, such as older age (AOR= (1.43, 1.72)), peers’ consumption of alcohol (AOR= (2.25, 4.84)), and parental drug problems (AOR= (1.28, 2.61)). Additionally, having peers who consumed marijuana (AOR= (2.02, 4.59)) also significantly increased the odds of nicotine use. Lower parental education (AOR Graduate Degree= (0.41, 0.87)) and unmarried parental status (AOR Married= (0.53, 0.99)) were associated with increased nicotine use.

Contrary to nicotine and alcohol, cannabis use was not associated with older age (AOR= (0.94, 1.29)). On the other hand, sex was significant, where male youth (AOR= (1.10, 2.07)) had increased odds of cannabis use. Peer alcohol (AOR= (1.16, 2.76)) and marijuana use (AOR= (12.56, 28.69)) were significantly associated with higher odds of cannabis use. Finally, parental drug problems (AOR= (1.57, 3.88)) were also significantly associated.

Compared to substance single use, multiple substance use had similar significant factors, such as older age (AOR= (2.05, 2.80)), peer consumption of alcohol (AOR= (2.64, 6.22)) and marijuana (AOR= (5.01, 11.47)), marriage status (AOR= (0.42, 0.90)), and parental substance use (AOR= (1.08, 3.76)). Additionally, White youth had increased odds compared to Black youth (AOR= (0.26, 0.75)), as it’s seen with alcohol use only. Other peer-related factors that showed strong associations where peers selling substances (AOR= (1.29, 2.95)) and peers consuming other substances (AOR= (1.26, 4.38)) were all significantly associated with increased odds of multiple substance use.

## DISCUSSION

Our study analyzed how demographic, family, and peer factors are associated with alcohol, nicotine, and cannabis use in children using data from the Adolescent Brain and Cognitive Development (ABCD) Study. Our findings highlight the significant role of peer behaviors, parental substance use, parental house rules, age, and race in influencing substance use among children as young as 9 to 13 years old. Additionally, our findings suggest that factor significance differs for each substance and for multiple substance use.

One of the primary findings is the strong association between peer substance use and children’s own use of alcohol, nicotine, cannabis, and multiple substances. Peer use of alcohol is significantly associated with using every type of substance, followed by peer use of marijuana, which increases the risk of children consuming nicotine, marijuana, or multiple substances during the year. This finding reinforces prior evidence that peer behaviors are among the most notable predictors of youth substance use.

Several parental factors were also significant. Parental drug use was consistently associated with higher odds of nicotine, cannabis, and multiple substance use. Additionally, parental permissiveness regarding alcohol use at home was strongly linked with alcohol use among children. Lower parental education and unmarried parental status were associated with higher odds of nicotine and multiple substance use. These findings emphasize the role of family environment and parental behaviors in shaping children’s risk for early substance use.

Age was an important demographic factor across substance categories. Older youth had higher odds of using alcohol, nicotine, and multiple substances, though interestingly, cannabis use was not significantly associated with age. Racial differences were also observed. White youth had a higher risk of alcohol use compared to other races but lower risk of cannabis use, compared to Blak youth, and higher risk of multiple substance use.

This study has several limitations. First, the data relies on self-reported substance use and peer behaviors, which may be subject to recall bias or social desirability bias. While the ABCD Study uses validated instruments, underreporting of substance use remains a concern, particularly among younger children. Second, the sample represents the current ABCD cohort and may not be generalizable to all populations, particularly as substance use patterns and social norms evolve over time. Third, we included select model predictors based on plausible relationship with substance use, where some potential risk factors were possibly overlooked.

Despite these limitations, this study offers important insights into early substance use in children. Our study shows that peer influence, parental substance use, and family dynamics are key factors associated with substance use in children. Additionally, relevant factors differ depending on the studied substance. These findings provide valuable information for the design of public health interventions aimed at reducing substance use initiation in youth populations. Early interventions targeting high-risk families and peer groups may reduce the initiation and progression of substance use in youth. Finally, continued longitudinal research with the ABCD cohort will be essential to understanding how these risk factors evolve as children transition into adolescence and young adulthood.

